# Impact of a medical-legal partnership program on readmissions to a family medicine inpatient service

**DOI:** 10.1101/2020.07.12.20152009

**Authors:** Emma James, Emily Samson, Megha Manek, Robert Robinson

**Affiliations:** Department of Obstetrics Gynecology, SIU School of Medicine, Springfield IL, USA; Department of Family Medicine, University of North Carolina School of Medicine, Chapel Hill, NC, USA; Department of Family and Community Medicine, SIU School of Medicine, Springfield IL, USA; Department of Internal Medicine, SIU School of Medicine, Springfield IL, USA

**Keywords:** medical legal partnership, hospital readmission, social determinants of health

## Abstract

**Background:** The Centers for Medicare and Medicaid Services (CMS) value-based reimbursement prioritizes health outcomes and population health in a way that emphasizes social determinants of health, access disparities, and prevention. Unmet legal assistance needs are an area of considerable interest because it can impact access to healthcare services and can be a social determinant of health. Medical-Legal Partnerships (MLP) are tools to link patients with unmet legal needs with lawyers and other legal services. The goal of these programs is to address legal issues that complicate the life and healthcare of patients. With the hypothesis that unmet legal needs were a risk factor for poor health outcomes, we studied the impact of unmet legal needs on the readmission rate to the SIU Family and Community Medicine inpatient service at Memorial Medical Center in Springfield, Illinois.

**Methods:** All adult patients discharged between January 2016 and March 2018 were retrospectively studied to determine if the need for legal services was a significant predictor of all-cause hospital readmission within 30 days of discharge. Patients in need of legal services were identified using an interview-based screening tool and were referred to an HRSA-grant funded MLP. The impact of unmet needs for legal services was compared to validated risk factors for hospital readmissions such as the LACE index and HOSPITAL score.

**Results:** Data from 2500 hospital discharges from the SIU FCM service were analyzed. The overall readmission rate was 27%. Univariate analysis showed patients who were readmitted were older (57 vs 53 years, p < 0.001), had longer hospital stays (4.69 vs 4.17 days, p < 0.014), had higher Charlson Comorbidity Index values (2.31 vs 1.57, p < 0.001), HOSPITAL scores (4.79 vs. 3.14, p < 0.001), LACE index values (10.13 vs. 8.33, p < 0.001) and were more likely to need legal services (12% vs. 3%, p < 0.001). Multivariate analysis showed the HOSPITAL score (OR 2.12, p<0.001), LACE index (OR 1.16, p<0.001), Charlson Comorbidity Index (OR 0.70, p<0.001), hospital length of stay (OR 0.85, p<0.001), congestive heart failure (OR 2.10, p<0.001), COPD (OR 1.85, p<0.001), diabetes with complications (OR 1.88, p<0.001), and a need for legal services (OR 2.21, p<0.001) to be independent predictors of an increased risk of hospital readmission.

**Conclusion:** Unmet legal needs identified in this study via an interview-based screening tool appear to be an independent risk factor for hospital readmission. The predicted risk of readmission (OR 2.21) compares favorably with validated HOSPITAL score (OR 2.12) and LACE index (OR 1.16) readmission risk assessment tools. These findings suggest that the presence of unaddressed legal issues may be a modifiable non-medical risk factor for hospital readmission. Further study of the impact of MLP programs on hospital readmission is necessary to clarify the implications of this social determinant of health on a critical value-based care measurement.

## Introduction

Hospital readmissions are common and expensive, with nearly 20% of Medicare patients being readmitted to a hospital within 30 days of discharge at an overall cost of nearly 20 billion US dollars per year (Jencks, Williams and Coleman 2009). Because of this high frequency and cost, hospital readmissions within 30 days of discharge are a target for health care cost savings in the Medicare Value Based Purchasing (VBP) program. The VBP aims to incentivize hospitals and health systems to reduce readmissions through reductions in payments to hospitals with higher than expected readmission rates (Centers for Medicare and Medicaid Services, 2016). Because of the VBP initiative, health care organizations are investing considerable resources into efforts to reduce hospital readmission.

The underlying risk factors for hospital readmission are diverse. Studies have identified age, race, having a regular health care provider, major surgery, medical comorbidities, length of hospital stay, previous admissions in the last year, failure to transfer important information to the outpatient setting, discharging patients too soon, the number of medications at discharge, and many other risk factors for hospital readmission within 30 days (Auerbach et al, 2016; Picker et al., 2015; Hasan et al, 2010; Silverstein et al., 2008).Medical Legal Partnerships (MLP) incorporate lawyers, usually from civil legal aid organizations, into the medical team to address patients’ unmet legal needs that may be affecting their health (National Center for Medical Legal Partnership, 2019). Unaddressed legal needs often include issues with income, health insurance, housing, employment, immigration status, and family stability (National Center for Medical Legal Partnership, 2019). It is estimated that that 60% of premature deaths in the USA are attributable to the social determinants of health – environmental exposure, social circumstances, and behavioral patterns (Schroeder, 2007). MLPs have the potential to improve health status by addressing unsafe housing, food insecurity, income for disabled individuals, orders of protection, and other unmet civil legal issues. One study found addressing the legal needs of high-need high-healthcare-use patients decreased inpatient and emergency department use by 50% and overall healthcare costs by 45% (Martin, Martin, Shultz, and Sandel 2015). A rural MLP program in Illinois demonstrated a 319% return on investment between 2007-2009 (Teufel et al, 2012).

The current investigation will explore if unaddressed legal needs, identified by the MLP screening tool, are significant predictors of hospital readmission. The performance of unmet legal needs as a risk factor for hospital readmission will be directly compared to the validated HOSPITAL and LACE readmission risk prediction tools. The HOSPITAL score and LACE index are useful predictors of readmission risk that are focused on the severity of illness and medical complexity.

## Materials and Methods

Patients discharged from Memorial Medical Center between January 2016 and March 2018 by the SIU-School of Medicine (SIU-SOM) Family and Community Medicine inpatient service were retrospectively studied to determine if the need for legal services was a significant predictor of any cause hospital readmission within 30 days of discharge. Patients who died while in the hospital, were transferred to another hospital, or left against medical advice were excluded from analysis. The study endpoint was all-cause hospital readmission within 30 days of discharge. Memorial Medical Center is university-affiliated not-for-profit tertiary care center located in Springfield, Illinois, USA. The SIU-SOM Family and Community Medicine inpatient service is the family medicine residency teaching service and is staffed by board certified or board eligible faculty. Most patients for this service are admitted via the hospital emergency department. The MLP is an HRSA grant funded program designed to provide patients with free legal assistance through the Land of Lincoln Legal Assistance Foundation (LOLLAF) to address social and civil legal issues that may have a negative impact on the patient. Patients of SIU-SOM Family and Community Medicine practice are eligible to receive legal assistance from a legal assistance lawyer who works full time for SIU patients. MLP incorporates legal services into the healthcare team of patients at SIU’s Center for Family and Community Medicine FQHC. Second-year medical students serve as inpatient screeners at Memorial Medical Center. Students screen patients using a questionnaire developed by the legal aid lawyers (Figure 1). All patients admitted to the family medicine inpatient service were eligible for screening. Common reasons for not being screened include patient declining screening, medical instability, and delirium. Patient screening occurred only on weekdays during the medical student academic year, from August to May. Patients who screened positive for possible legal issues signed HIPAA release for necessary contact information and this information was faxed to the Land of Lincoln Legal Aid office. Follow up appointments were made after further discussion with the Intake Specialist at LOLLAF. De-identified data on hospital readmissions, length of stay, diagnosis-related group (DRG), International Classification of Disease (ICD) diagnosis codes, age, gender, and the other variables in the HOSPITAL score (Table 1) and LACE index (Table 2) were extracted from the electronic health record. Patients screening positive for unaddressed legal needs were compared to the remainder of the family medicine inpatient service discharges. The study hospital does not have an admitting service for oncology patients, so patients with any oncology-related ICD codes were classified as being discharged from an oncology service for the purposes of calculating a HOSPITAL score. This reflects local practice patterns and accounts for the increased risk of readmission found in oncology patients. As a single center study, readmissions at other hospitals will not be detected. This study protocol was reviewed by the Springfield Committee for Research Involving Human Subjects, the local institutional review board (IRB). The IRB determined this study did not meet the criteria for research involving human subjects defined by 45 CFR 46.101 and 45 CFR 46.102.

**Figure 1.**
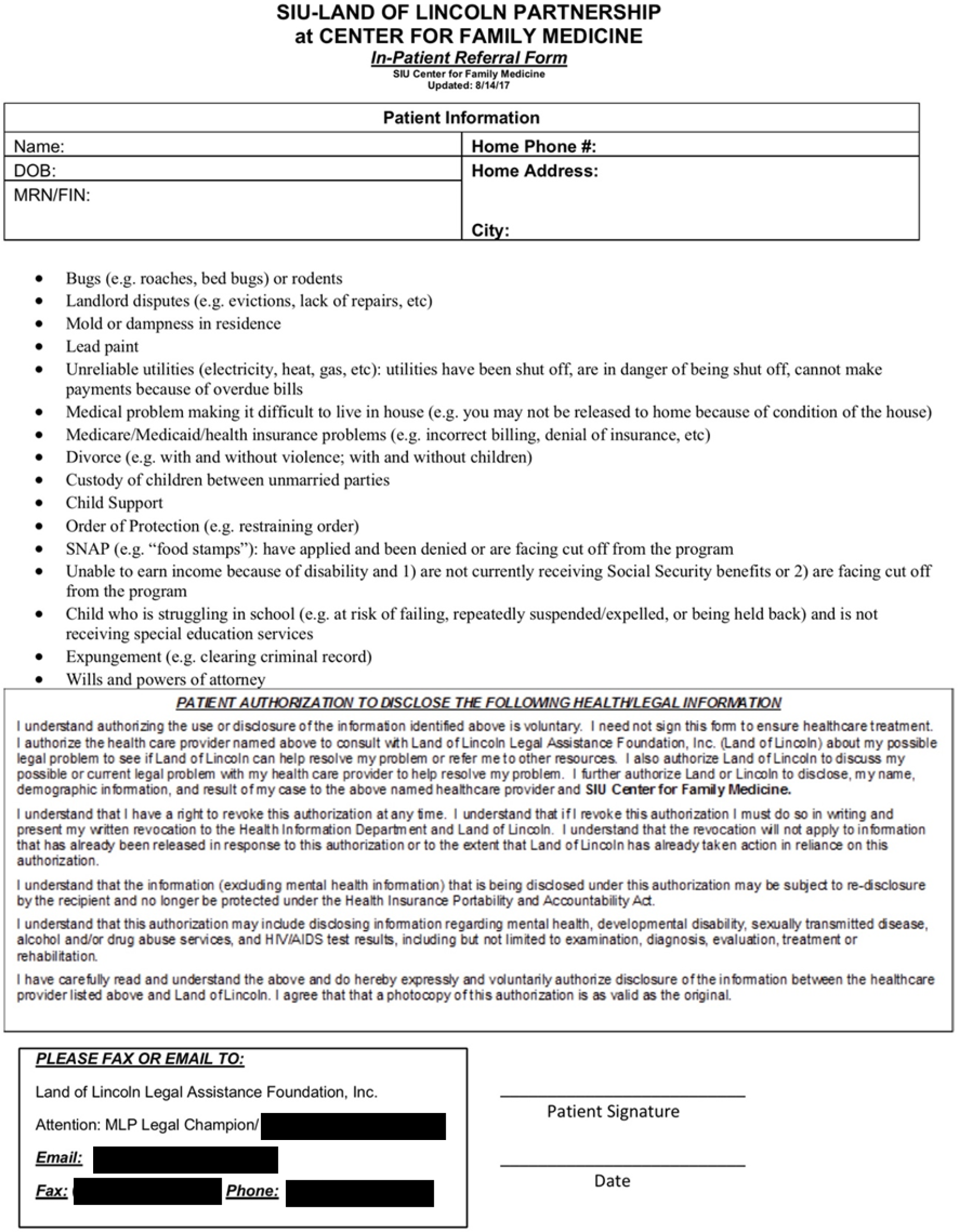
Legal Needs Screening Survey

**Table 1.** The HOSPITAL score

| Attribute | Points if Positive |
| --- | --- |
| Low hemoglobin at discharge (<12 g/dL) | 1 |
| Discharge from an Oncology service | 2 |
| Low sodium level at discharge (<135 mEq/L) | 1 |
| Procedure during hospital stay (ICD10 Coded) | 1 |
| Index admission type urgent or emergent | 1 |
| Number of hospital admissions during the previous year |  |
| 0-1 | 0 |
| 2-5 | 2 |
| >5 | 5 |
| Length of stay ≥ 5 days | 2 |

**Table 2.**
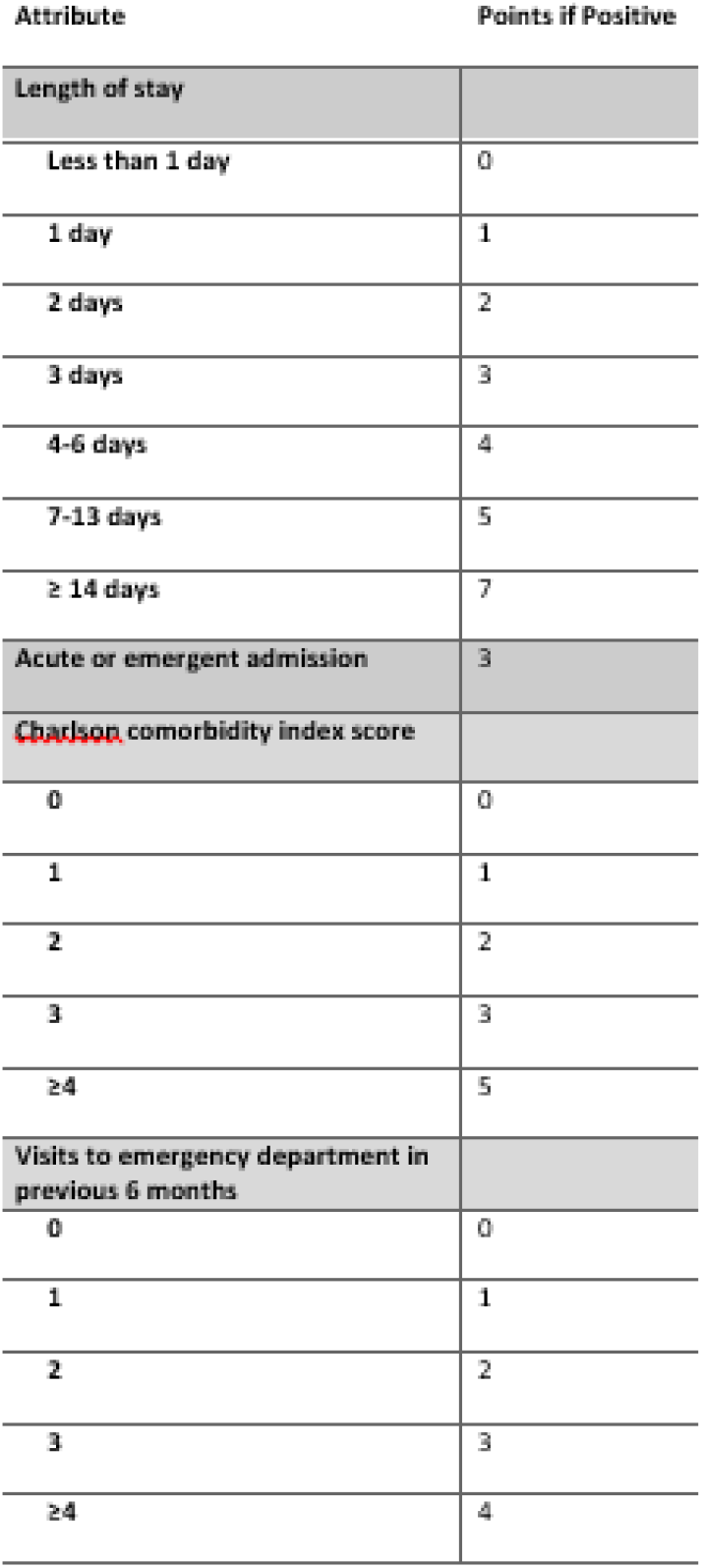
The LACE Index

### Statistical analysis

The need for legal services through the MLP program was investigated as a predictor of any cause hospital readmission within 30 days. A HOSPITAL score, LACE index, and Charlson score were calculated for each admission. The Pearson chi-squared or Fisher’s exact test were used to evaluate qualitative variables. The Mann–Whitney U or Kruskal–Wallis tests were used to evaluate quantitative variables. Variables from univariate analysis with a p-value of 0.05 or less were evaluated using multivariate logistic regression with backward likelihood ratio based method. Two sided P-values < 0.05 were considered significant. Statistical analysis was performed using SPSS version 22 (SPSS Inc., Chicago, IL, USA).

## Results

The SIU-SOM Family and Community Medicine inpatient service had 2645 discharges recorded between January 2016 and March 2018. Inclusion criteria were met in 2500 discharges, of which 678 were readmitted within 30 days (Figure 2). Of those 2500, 8.4% (210) were screened by the MLP program. Legal services were provided to 5.3% (134) of the patients.

**Figure 2.**
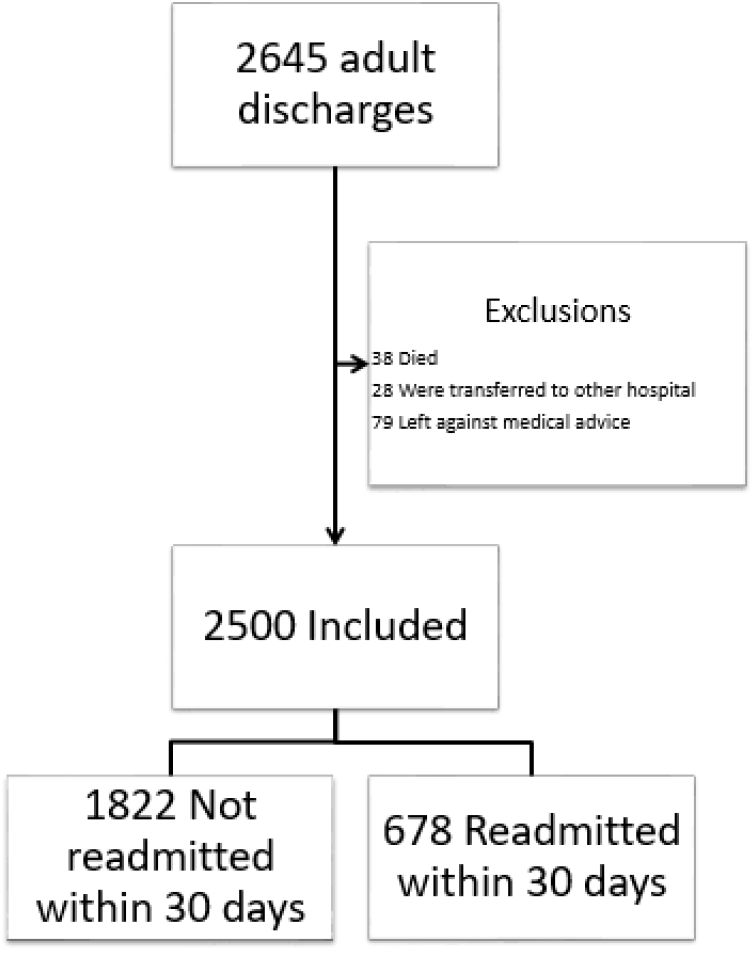
Study Flow Diagram

Patients who were readmitted were older (57 vs 53 years, p < 0.001), had longer hospital stays (4.69 vs 4.17 days, p < 0.014), and had higher Charlson Comorbidity Index values (2.31 vs 1.57, p < 0.001). These patients had higher HOSPITAL scores (4.79 vs. 3.14, p < 0.001) and LACE index values (10.13 vs. 8.33, p < 0.001), indicating a higher risk of hospital readmission. Higher rates of myocardial infarction, CHF, peripheral vascular disease, cerebrovascular disease, COPD, diabetes with complications, advanced renal disease, and liver disease were seen in the group readmitted to the hospital (Table 3). Patients who had screened positive for unresolved legal issues and received linkage to legal services through the MLP program were more likely to be readmitted (12% vs. 3%, p < 0.001).

**Table 3.**
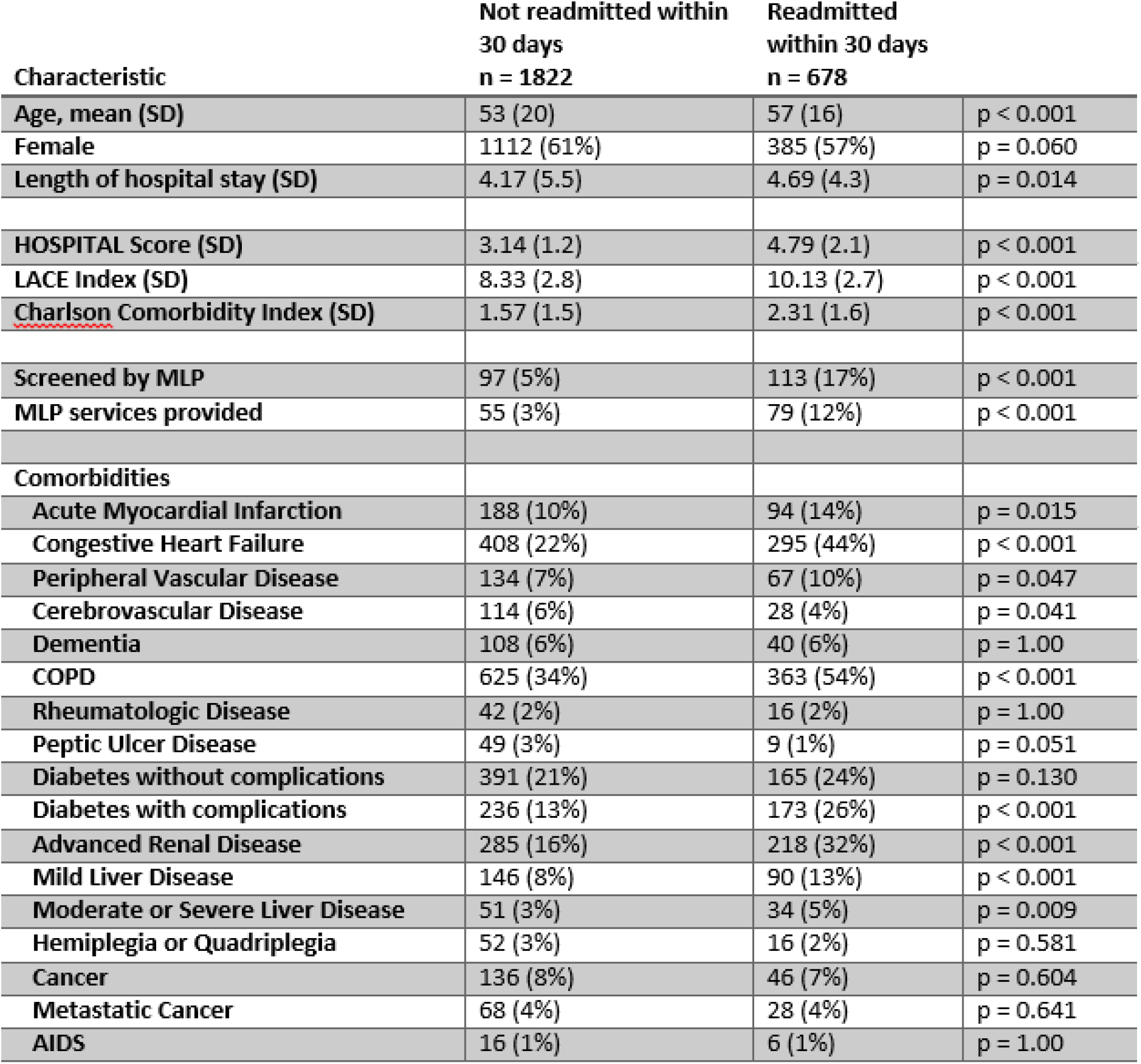
Baseline characteristics of the study population by 30 day readmission status

| Characteristic | Not readmitted within<br>30 days<br>n = 1822 | Readmitted<br>within 30 days<br>n = 678 |  |
| --- | --- | --- | --- |
| Age, mean (SD) | 53 (20) | 57 (16) | p < 0.001 |
| Female | 1112 (61%) | 385 (57%) | p = 0.060 |
| Length of hospital stay (SD) | 4.17 (5.5) | 4.69 (4.3) | p = 0.014 |
| HOSPITAL Score (SD) | 3.14 (1.2) | 4.79 (2.1) | p < 0.001 |
| LACE Index (SD) | 8.33 (2.8) | 10.13 (2.7) | p < 0.001 |
| Charlson Comorbidity Index (SD) | 1.57 (1.5) | 2.31 (1.6) | p < 0.001 |
| Screened by MLP | 97 (5%) | 113 (17%) | p < 0.001 |
| MLP services provided | 55 (3%) | 79 (12%) | p < 0.001 |
| Comorbidities |  |  |  |
| Acute Myocardial Infarction | 188 (10%) | 94 (14%) | p = 0.015 |
| Congestive Heart Failure | 408 (22%) | 295 (44%) | p < 0.001 |
| Peripheral Vascular Disease | 134 (7%) | 67 (10%) | p = 0.047 |
| Cerebrovascular Disease | 114 (6%) | 28 (4%) | p = 0.041 |
| Dementia | 108 (6%) | 40 (6%) | p = 1.00 |
| COPD | 625 (34%) | 363 (54%) | p < 0.001 |
| Rheumatologic Disease | 42 (2%) | 16 (2%) | p = 1.00 |
| Peptic Ulcer Disease | 49 (3%) | 9 (1%) | p = 0.051 |
| Diabetes without complications | 391 (21%) | 165 (24%) | p = 0.130 |
| Diabetes with complications | 236 (13%) | 173 (26%) | p < 0.001 |
| Advanced Renal Disease | 285 (16%) | 218 (32%) | p < 0.001 |
| Mild Liver Disease | 146 (8%) | 90 (13%) | p < 0.001 |
| Moderate or Severe Liver Disease | 51 (3%) | 34 (5%) | p = 0.009 |
| Hemiplegia or Quadriplegia | 52 (3%) | 16 (2%) | p = 0.581 |
| Cancer | 136 (8%) | 46 (7%) | p = 0.604 |
| Metastatic Cancer | 68 (4%) | 28 (4%) | p = 0.641 |
| AIDS | 16 (1%) | 6 (1%) | p = 1.00 |

Multivariate analysis showed the HOSPITAL score (OR 2.12, p<0.001), LACE index (OR 1.16, p<0.001), Charlson Comorbidity Index (OR 0.70, p<0.001), hospital length of stay (OR 0.85, p<0.001), congestive heart failure (OR 2.10, p<0.001), COPD (OR 1.85, p<0.001), diabetes with complications (OR 1.88, p<0.001), and a need for legal services (OR 2.21, p<0.001) to be independent predictors of an increased risk of hospital readmission. (Table 4).

**Table 4.** Multivariate logistic regression of potential risk factors for hospital readmission within 30 days of discharge.

|  | Odds Ratio (95% CI) | P value |
| --- | --- | --- |
| **HOSPITAL Score** | 2.12 (1.94 – 2.31) | <0.001 |
| **LACE Index** | 1.16 (1.08 – 1.24) | <0.001 |
| **Charlson Comorbidity Index** | 0.70 (0.60 – 0.81) | <0.001 |
| **MLP Assisted** | 2.21 (1.42 – 3.44) | <0.001 |
| **Medical comorbidities** |  |  |
| CHF | 2.10 (1.56 – 2.84) | <0.001 |
| COPD | 1.85 (1.44 – 2.38) | <0.001 |
| Diabetes with complications | 1.88 (1.37 – 2.55) | <0.001 |
| Mild liver disease | 1.41 (0.99 – 2.01) | 0.058 |
| **Length of hospital stay** | 0.853 (0.82 – 0.89) | <0.001 |

A receiver operating characteristic (ROC) evaluation (Figure 3) showed that the HOSPITAL score and LACE index had good discrimination for hospital readmission in this population. The HOSPITAL score had a C statistic of 0.74 (95% CI 0.72 – 0.76) with a p-value of less than 0.001. The LACE index had a C statistic of 0.68 (95% CI 0.66 – 0.70) with a p-value of less than 0.001. The need for MLP services had a C statistic of 0.54 (95% CI 0.52 – 0.57) with a p-value of 0.001.

**Figure 3.**
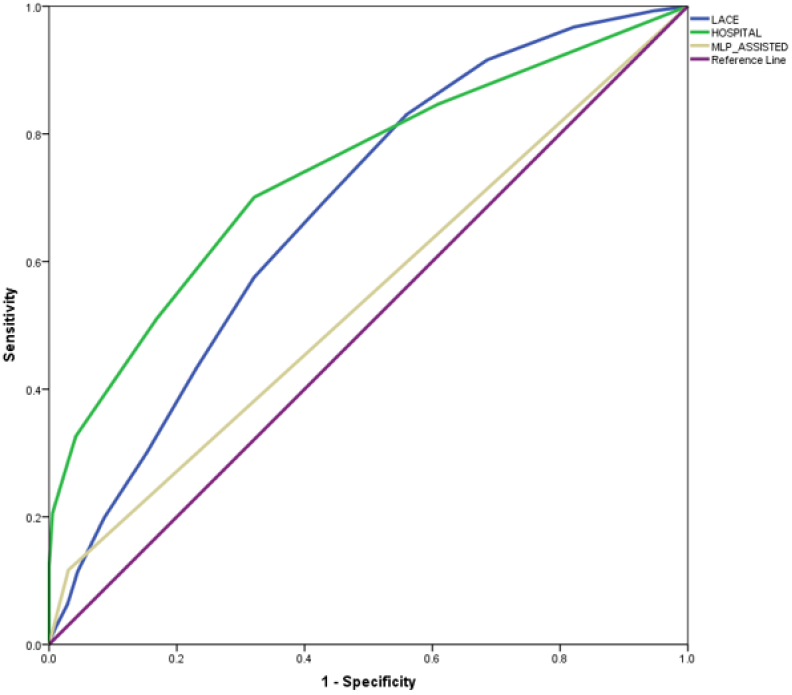
Receiver operating characteristic curves for the HOSPITAL score, LACE index, and the need for services from the MLP

## Discussion

The results indicate that many factors predict readmission in the study population, including older age, longer hospital stays, higher Charlson Comorbidity Index values, higher HOSPITAL scores, and higher LACE indices. Linkage to legal services for unmet legal need was also a significant predictor of hospital readmission. The receiver operating characteristics show the HOSPITAL and LACE indices to be strong models of predicting 30-day readmission. Patients that screened positive for unmet legal issues and were subsequently linked to MLP legal aid lawyers also achieved significance as a model of predicting readmission, albeit a much weaker model. The importance of this finding is that legal need, unlike many predictors of readmission such as age and comorbidities, is subject to intervention.

Multivariate analysis showed unmet legal needs (OR 2.21) compared favorably with the validated HOSPITAL score (OR 2.12) and the high risk for readmission medical problems of congestive heart failure (OR 2.10) and COPD (1.85). Unmet legal need differs substantially from these other risk factors because legal services have the potential to eliminate this risk factor, unlike interventions for COPD or CHF that manage but do not resolve the disease process. Unmet legal needs appear to be a new member of a short list of potentially modifiable risk factors for hospital readmission which include polypharmacy,(Picker, Heard, Bailey et al. 2015) malnutrition (Sharma, Miller, Kaambwa et al. 2017; Budzynki, Tojek, Chzernak et al. 2016; Sanchez-Rodriguez, Annweiler, Ronquillo-Moreno et al. 2019), and substance abuse (Gerke, Agley, Wilson et al. 2018).

Patients in the current investigation were connected with legal aid lawyers but the vast majority had not received resolution of their legal issue by the end of the 30-day readmission timeframe. On average, it takes one to two weeks after referral to schedule an initial meeting with the legal aid lawyer. The potential positive effects of the lawyer’s intervention are unlikely to be captured in a 30-day readmission. The differences seen during this time frame are more likely a result of the presence of unmet legal needs regarding food, income, insurance, or housing. This study is retrospective, single center, and did not investigate if the unmet legal needs directly led to hospital readmission.

## Conclusions

Patients that screened positive for unmet legal issues and were subsequently linked to MLP legal aid lawyers were significantly more likely to be readmitted to the hospital within 30 days. This implies that unmet legal needs are an independent risk factor for hospital readmission, and therefore a potential point for intervention and reduction of 30-day readmission. Unaddressed legal issues may be a modifiable non-medical risk factor for readmission.

The vast majority of patients do not reach resolution of their legal issue within 30 days, and most take one to two weeks to schedule an appointment with the lawyer. Analysis of the effect of the legal intervention would likely be seen after 30 days. When patients reach resolution of their legal issues through the help of the MLP lawyers their readmission rate may return to baseline, potentially decreasing the healthcare system resources spent on preventable hospital readmission. Future analysis will focus on the effect of legal aid intervention on the modification of readmission risk.

## Data Availability

No data is available for reuse.

## AUTHOR CONTRIBUTIONS

Study design EJ, ES, RR Supervision of MLP program - EJ, ES, MM Data acquisition - EJ, ES, RR Statistical analysis - RR Writing, original draft - EJ, ES Writing, review and editing - EJ, ES, MM, RR

